# Paediatrics Health Outcomes in Sub-Saharan Africa: A Multi-Country Assessment of Antecedence of Childhood Vaccination Decision-Making

**DOI:** 10.64898/2026.03.12.26348244

**Authors:** Gbadebo Collins Adeyanju, Lars Korn

**Author notes:** **Corresponding Author:** Gbadebo Collins Adeyanju Center for Empirical Research in Economics and Behavioral Science (CEREB) University of Erfurt, Erfurt, Germany.

## Abstract

**Background:** Vaccination is a cost-effective intervention preventing causes of 48% of deaths among children Under-5 in sub-Saharan Africa. However, one in five African children still has not completed basic vaccination, and over six million children have not received a single dose of the Diphtheria, Tetanus and Pertussis vaccine, resulting in over half a million deaths annually. This study aims to understand the key factors influencing pediatrics health (vaccination) decision-making in sub-Saharan Africa.

**Methods:** A cross-sectional design using a multi-stage stratified sampling approach was used. Data were collected from 2,451 households with children Under-5 in three countries and analyzed using R. Correlation analysis was used to understand the associations between variables, while regression analysis was used to control for covariates and identify influences on one another.

**Results:** The findings show that beliefs in gender disparity, misinformation, and masculinity can undermine childhood vaccination outcomes. A child’s gender (*r* = –0.33–0.40, *p* < 0.01) and misinformation (*r* = –0.38–0.54, *p* < 0.01) impact vaccination intentions and behavior. Meanwhile, positive attitudes (*r* = 0.36–0.49, *p* < 0.01), trust (r = 0.17–0.34, *p* < 0.01) and peer influence (*r* = 0.27–0.33, *p* < 0.01) significantly improve uptake. However, regressions show that male caregivers had weaker attitudes (β = –0.31, *p* < 0.001), stronger beliefs about misinformation (β = 0.33, *p* < 0.001) and lower vaccination intentions (β = –0.12, *p* < 0.001). Country comparisons reveal that Kenyan and Malawian children are 3.6 (OR = 3.64, *p* < 0.001) and 13 (OR = 13.13, *p* < 0.001) times more likely to be vaccinated than Nigerian children, respectively. Furthermore, masculinity had a significant effect on men, Muslims and polygamous households.

**Conclusion:** To address low vaccination uptake in sub-Saharan Africa, context-sensitive strategies are required that incorporate gender norms, counter misinformation, and engage fathers.

## INTRODUCTION

Vaccination is one of the most significant scientific achievements in public health, particularly in combatting vaccine-preventable diseases (VPDs) in limited-resource settings. Globally, around 85% of infants (116 million) have received three doses of the Diphtheria-Tetanus-Pertussis (DTP3) vaccine, also known as the basic vaccination, as well as one dose of the Menin-gococcal conjugate vaccine (MCV) [1]. Over 123 countries have reported achieving DTP3 coverage of at least 90%, which is crucial in protecting children against preventable infectious diseases in childhood [2]. Currently, vaccines prevent over 4 million childhood deaths annually, and this figure could rise to over 50 million by 2030 [3,4]. Vaccines are an effective firstline defence strategy against VPDs, preventing deaths and mitigating the occurrence of disabilities such as neurodevelopmental disorders, hearing loss or deafness, meningitis, blindness, and impaired mobility. Today, vaccines play a significant role in preventing not only infectious diseases, but also other diseases such as cancers and, more recently, Malaria [5–7]. Furthermore, vaccination brings children and families into interaction with health systems, providing access to other essential healthcare services [8].

Through vaccination, the prevalence of vaccine-preventable child deaths has declined, especially among children Under-5, falling from 12.6 million in 1990 to 5.4 million in 2020 [9]. During this period, the number of Under-5 deaths globally decreased by about 60%, from an average of 93 to 37 per 1,000 live births [10,11]. Vaccination prevents the illnesses and infections that together account for 48% of deaths among children Under-5 [13]. While this is not yet at the desired level, it represents remarkable growth compared to previous years. However, although there has been unprecedented progress occasioned by increased vaccination, the rate of decline in Under-5 mortality slowed by half in the first half of the Sustainable Development Goals (SDGs) era (2015 – 2022) compared to the Millennium Development Goals (MDGs) era (2000–2015) [11]. Global DTP3 coverage remains well below the target of 90% [13].

In sub-Saharan Africa (SSA), immunization has become the cornerstone for reducing morbidity and mortality from VPDs, averting 154 million deaths, including 146 million among children Under-5 [14]. Childhood vaccination in the region provides a low-cost protection against preventable diseases, making it critical for achieving SDG 3, which aims to reduce Under-5 mortality to less than 25 per 1,000 live births by 2030 [15].

However, despite these achievements and prospects, immunization coverage has stagnated or even declined in many countries, particularly in the SSA region, where access issues and behavioral challenges remain significant [16–20]. While Under-5 mortality worldwide has fallen to 37 deaths per 1,000 live births, SSA countries still to have the highest mortality rates, at 74 deaths per 1,000 live births – 14 times higher than in Europe and North America [10]. In the SSA region, one in five children has not received basic vaccines, meaning over 30 million children Under-5 are at risk of VPDs [8]. The majority of the 20 million children worldwide who have not completed basic vaccination live in the SSA, with over 75% (15.7 million) being zero-dose or unvaccinated children, i.e., having not received a single dose of the life-saving DTP vaccine [5,21]. Furthermore, in 2024, over 20 million children have not received their first routine dose of the measles vaccine, a worsening situation compared to 19.3 million in 2019 [1]. Almost half of the nearly four million child deaths worldwide from VPDs occur in just five countries, three of which are in the SSA region [22].

The country-specific phenomenon is no different to that seen across the SSA region. In fact, some countries are worse. For example, Nigeria accounts for 30% of the world’s unvaccinated children Under-5 and is a major contributor to the Under-5 mortality, with 132 deaths per 1,000 live births [22]. I.e., one in thirteen Nigerian children dies before their first birthday. Nigeria has 2.1 million zero-dose children, representing the largest population of unvaccinated children worldwide [12]. Demographically, Nigeria has 36 million children Under-5, representing over 16% of its total population - one of the highest Under-5 populations in the world [12]. This implies that Nigeria bears the greatest proportion of the global burden of zero-dose and underimmunized children, yet immunization coverage is estimated to be only 54% [9,23]. Even worse, full immunization coverage in all its Northern states is below 40% [24]. Similarly, although Malawi has maintained high immunization coverage (>80%) for various antigens for decades, the percentage of fully immunized children has recently declined to 76%, and with a 4% dropout rate in basic vaccination threatening the sustainability of current uptake [2,6,25]. Although Kenya’s basic vaccination coverage exceeds 80%, it faces tremendous challenges with regards to zero-dose children, who represent about 13% of the population, with rates reaching 33.8% and 35.1% in Wajir and Garissa counties, respectively [26]. Nevertheless, one in ten Kenyan children remains unvaccinated, and two in three Kenyan girls have not received at least one dose of the Human Papillomavirus (HPV) vaccine [27].

Consequently, prospects for improving childhood vaccination are under serious threat due to the recent decline in vaccination coverage, resulting in increased outbreaks of VPDs in the region [22]. This is largely attributable to vaccine hesitancy, among other factors [5,6,22,27,28]. Driven by misinformation, conspiracy theories, inadequate access to vaccination services and weaknesses in the health system, among others, vaccine hesitancy is threatening to undermine the significant gains made in achieving basic vaccine uptake in countries across the SSA [5,6,22,28].

Therefore, it is important to assess the processes surrounding caregivers’ vaccination behavior and the underlying motivations behind decision-making, given the dearth of in-depth research on behavioral mechanisms in SSA vaccination. Even where successes have been recorded in terms of vaccination uptake in the SSA, threats to the sustainability of these gains have emerged in recent years due to various factors, including the roles of misinformation, the gender of the child, masculinity or attitudes of fathers toward vaccination, and trust [15–17,29,30]. Despite the growing body of research on vaccine hesitancy, little is known about how masculinity norms, paternal authority, gender disparity, and the weaponization of inaccurate information shape vaccination decision-making in SSA households. Consequently, unless the sociodemographic, social, and psychological influences associated with childhood vaccination behavior and decision-making are properly understood and addressed through targeted interventions, low vaccination demand and the resultant infant mortality and morbidity will persist [17,30].

Barriers to childhood vaccination coverage are inherent in the attitudes of caregivers, health systems and vaccination service providers. However, the decision-making barriers faced by caregivers appear to be more significant and consistent than those faced by providers and health systems [15]. Existing research has largely focused on health system constraints and access-related barriers, paying comparatively little attention to the behavioral and psychosocial mechanisms that shape caregivers’ vaccination decisions in SSA contexts. In particular, empirical evidence remains sparse on how gender norms, masculinity, paternal authority, household power dynamics, misinformation and trust interact to influence vaccination intentions and behaviors within households.

The Theory of Planned Behavior (TPB) provides a useful framework for understanding vaccination decisions. It posits that an individual’s intention to perform a given behavior (such as whether or not to vaccinate one’s child) is shaped by three key determinants: personal attitudes towards the behavior (influenced by knowledge, attitudes and prejudices); subjective norms (e.g., perception of other people’s attitudes; community norms regarding health-related gender disparity; and shared perceptions of masculinity); and perceived behavioral control (the perceived ease or difficulty of performing the behavior, often shaped by access barriers or misinformation) [31,32]. This theoretical perspective aligns closely with the rationale of the study. The TPB assumes that vaccination decisions are not random or purely habitual but rather result from reasoning processes that are sometimes misinformed yet still informed. Applying this framework to the context of SSA reveals significant behavioral evidence gaps. While a range of structural and contextual factors contributing to low vaccination uptake have been identified, far less is known about how caregivers decide in SSA contexts, given the behavioral science research bias towards samples from the Global North. In particular, the psychosocial and normative mechanisms shaping childhood vaccination decisions in SSA are poorly understood. This including the influence of gender norms, the perceived role of fathers, community rumor networks, and levels of trust. This study therefore aims to address these gaps by examining the motivational and cognitive processes underlying parental vaccination behavior in SSA.

Based on this theoretical foundation, the study examines childhood vaccination in SSA through the lens of caregivers’ decision-making processes. Specifically, it seeks to understand how gender norms, masculinity, misinformation household power dynamics drive or hinder vaccination intention and behavior. Grounded in the theoretical assumptions of the TPB, the study hypothesizes that several psychosocial variables such as the child’s gender, fathers’ attitudes, notions of masculinity, trust, misinformation, and religious beliefs are likely to be negatively associated with vaccination intention and behavior. Conversely, higher perceived knowledge about vaccination is expected to positively predict both vaccination intention and behavior.

In addition, the study considers demographic characteristics, such as caregivers’ age, education level, household income, and country’s context, as potential predictors of vaccination decision-making. Understanding how these structural and psychosocial factors interact will help clarify the mechanisms underlying persistently low vaccination uptake in the SSA region.

## METHODS

### Study and sampling design

The study employed a cross-sectional design with a multi-stage, stratified sampling approach [33]. Data were collected in three countries: Nigeria, Kenya and Malawi. A structured questionnaire (see Supplementary File 1) was administered to eligible households with parents or legal guardians (hereafter referred to as caregivers) of children Under-5. In each country, study sites were selected to reflect the major geopolitical and sociocultural regions. Within these strata, communities were randomly selected and, within each community, eligible households were systematically sampled. In the final stage, one caregiver per household was invited to participate; typically, this was the head of the household or the primary decision-maker for the child’s health. A total of 2,451 caregivers participated. The overall sample was designed to capture national diversity and ensure representativeness across regions within each country. The sampling procedure is summarized in Figure 1.

**Figure 1:**
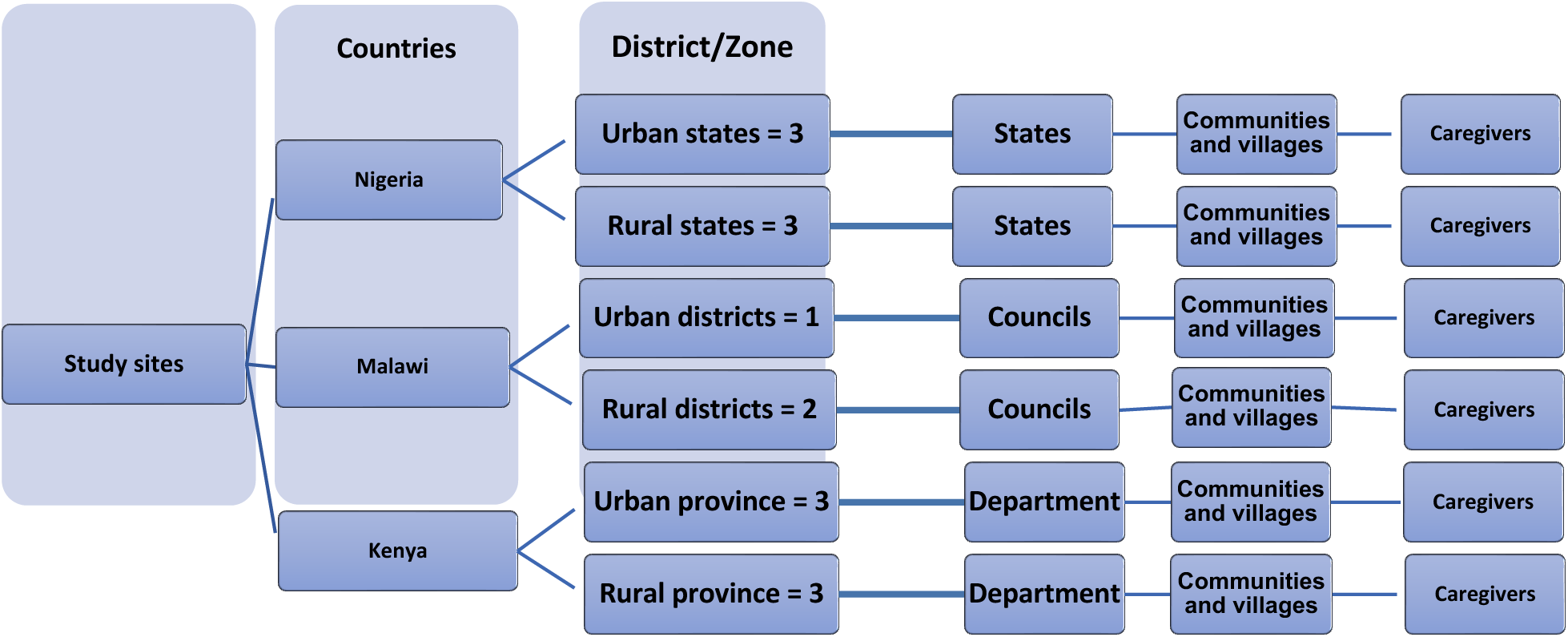
**Study sample plan**

### Study settings and participant characteristics

The participants were representative of the population in terms of age, gender, socioeconomic status and regional distribution. In Nigeria, one state was randomly selected from each of the six geopolitical zones: Sokoto, Kogi, Adamawa, Enugu, Osun, and Delta. The same sampling strategy was applied in Kenya, where participants were drawn from the provinces of Nairobi, Central, Rift Valley, Eastern, Coast, Nyanza, Western and North-Eastern. In Malawi, data were collected from the districts of Mzimba, Lilongwe and Blantyre. The targeted participants were caregivers of children Under-5 who were heads of household (HHs). As illustrated in Figure 2, the three study sites represent key sub-regions within the SSA region, with Nigeria, Kenya and Malawi reflecting West, East and Southern Africa respectively.

**Figure 2:**
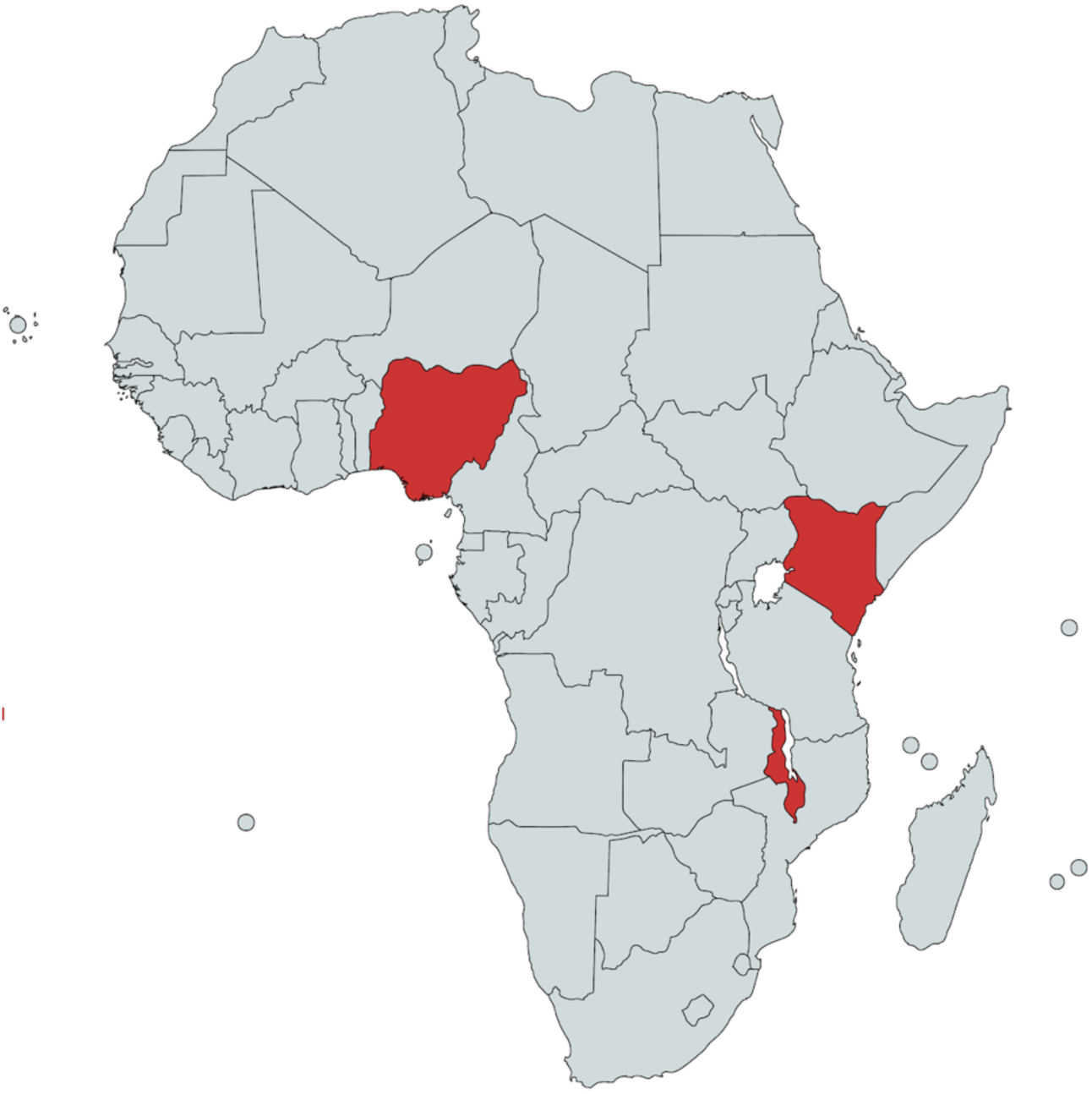
**Study Settings**

## Measures

### Main variables

*Intention to vaccinate*: This was assessed using a single item: “I will not vaccinate my child when the next vaccination appointment is due.” Responses were recorded on a five-point Likert scale ranging from *strongly disagree* to *strongly agree*. For the analysis, the item was reverse-coded, meaning that higher values indicated a greater intention to vaccinate the child. *Vaccination behavior:* Vaccination behavior was measured using one item: “Of the three doses of the combined DTP3 or Pentavalent vaccine, how many has your child received according to their age?” Response options were *none*, *some*, or *all*. For the analysis, the responses were dichotomized: participants who selected *none* or partial were coded as being consistent with the WHO and national immunization benchmarks. Behaviorally, full completion demonstrates sustained vaccination intention, whereas incomplete uptake indicates disruption to this process, potentially shaped by similar psychosocial barriers as non-vaccinated (0), reflecting vaccine hesitancy, while those who selected *all* were coded as vaccinated (1). *Gender disparity:* Gender disparity was measured with one item: “I think immunization is more important for girls than boys.” Responses were captured on a five-point Likert scale ranging from *strongly disagree* to *strongly agree*. *Misinformation:* Misinformation was assessed using three items (e.g., “Some vaccines generally contain chemicals that are harmful to children”), which were rated on a five-point Likert scale ranging from *strongly disagree* to *strongly agree*. A mean score was computed for analysis. The scale showed good reliability (Cronbach’s α = .731). *Masculinity:* Masculinity was measured using one central construct: “The final decision on our child’s immunization rests with the father.” This item was rated on a five-point Likert scale ranging from *strongly disagree* to *strongly agree*.

### Additional variables

*Vaccination attitude:* Attitudes towards vaccination were measured using three items: “How important do you think vaccines are for your child’s health?”, “Nigeria has recommended vaccines for children. How important is it for your child to receive these vaccines?”, and “I want my child to receive all childhood immunizations.” The first two items were rated on a four-point Likert-type scale ranging from *not important at all* to *very important*, while the third item was rated on a five-point Likert scale ranging from *strongly disagree* to *strongly agree*. These items were then combined to create composite measure representing general attitudes towards vaccination (Cronbach’s α = .726). *Trust:* Trust in sources of vaccination information was assessed using five items (e.g., “I trust the vaccination information I receive from family and friends”). Responses were given on a five-point Likert scale ranging from *strongly disagree* to *strongly agree*. A mean score was computed, and the scale demonstrated moderate reliability (Cronbach’s α = .642). *Perceived knowledge:* Perceived knowledge about childhood vaccination was measured using one item: “I think immunization prevents all childhood diseases.” Responses were rated on a five-point Likert scale ranging from *strongly disagree* to *strongly agree*. *Religious beliefs:* Religious beliefs concerning vaccination were assessed using one item: “Vaccination does not go well with my religious beliefs.” Responses were given on a five-point Likert scale ranging from *strongly disagree* to *strongly agree*. *Sources of vaccination information:* Participants were asked to identify their most influential source of vaccination information using the following item: “I get my most important vaccination information from….” Response options included social media, radio or television, family and friends, church or mosque, and healthcare workers (HCWs).

### Data analysis

First, descriptive statistics and bivariate correlations were computed to examine the relationships among the study variables. Both Pearson’s product-moment and Spearman’s rank correlations [34,35] were reported to provide an overview of linear and monotonic associations. Linear regression analyses [36] were then conducted to examine the predictors of gender disparity, masculinity, misinformation and intention to vaccinate. In addition, logistic regression analyses [37] were used to assess the effects of psychosocial factors on caregivers’ actual vaccination behavior. Demographic variables were included as control variables in all regression models to obtain a more accurate understanding of the relationships between psychological predictors and vaccination outcomes.

An alpha level of 0.05 was applied to all statistical tests. As participation in the survey was voluntary, missing data occurred across both dependent and independent variables, leading to variation in sample sizes between analyses. Missing data were handled using listwise deletion. This involved excluding cases with incomplete values for the variables included in each analysis.

## RESULTS

### Associations between variables

Pearson’s and Spearman’s correlations were computed to examine the relationships between vaccination outcomes and key psychosocial variables (see Supplementary File 2). As hypothesized, gender disparity was significantly and negatively correlated with both vaccination intention (*r* = –0.33, *p* < 0.01) and vaccination behavior (*r* = –0.21, *p* < 0.01). Caregivers who endorsed stronger gendered norms/gender disparity reported less favorable attitudes towards vaccination (*r* = –0.34, *p* < 0.01). Misinformation showed the strongest negative association with vaccination intention (*r* = –0.54, *p* < 0.01) and was also related to lower vaccination behavior (*r* = –0.29, *p* < 0.01) and less positive attitudes (*r* = –0.53, *p* < 0.01). Conversely, favorable attitudes towards vaccination were positively associated with both intention (*r* = 0.49, *p* < 0.01) and behavior (*r* = 0.36, *p* < 0.01), which supports the theoretical assumptions of the TPB. Trust in information sources was also positively related to attitudes (*r* = 0.34, *p* < 0.01) and intention (*r* = 0.17, *p* < 0.01) as well as vaccination behavior (*r* = 0.16, *p* < 0.01), suggesting that greater confidence in information channels promotes vaccine acceptance. Similarly, subjective knowledge and peer influence were positively associated with vaccination intention and behavior, though these associations were moderate in strength. Masculinity exhibited negative correlations with vaccination intention (*r* = –0.35, *p* < 0.001) and behavior (*r* = –0.17, *p* < 0.001) and was positively corelated with misinformation (*r* = 0.33, *p* < 0.001) and rumor-related beliefs (*r* = 0.19–0.28, *ps* < 0.001). This is highlighting the influence of patriarchal decision-making norms on vaccine hesitancy. Among the demographic factors examined, male caregivers reported significantly lower vaccination intention (*r* = –0.13, *p* < 0.01) and behavior (*r* = –0.13, *p* < 0.01), while higher household income was associated with greater intention (*r* = 0.22, *p* < 0.01) and behavior (*r* = 0.15, *p* < 0.01). Parental education level showed no meaningful association with vaccination outcomes.

### Primary outputs

#### Vaccination intention

A linear model estimated using ordinary least squares was applied to predict the intention to vaccinate a child, with psychological and social factors as main predictors, while demographic variables were included as controls (see Supplementary File 3 for details). The model explained a substantial proportion of the variance (adj. R² = 0.32, *p* < 0.001). The results showed that masculinity (β = –0.12, *p* < 0.001), misinformation (β = –0.29, *p* < 0.001), religious beliefs (β = –0.17, *p* < 0.001), and, counterintuitively, subjective knowledge of vaccination (β = –0.08, *p* < 0.001) were all negatively related to the intention to vaccinate a child. In contrast, vaccination attitudes were positively associated with vaccination intentions (β = 0.11, *p* < 0.001). Gender disparity (β = –0.02, *p* = 0.376) and trust in media sources (β = 0.03, *p* = 0.122) were not significant predictors. Meanwhile, compared to Nigerians, Kenyans (β = 0.11, 95% CI [2.75e-03, 0.23], p = 0.045) and Malawians (β = 0.25, 95% CI [0.07, 0.42], p = 0.005) had higher vaccination intentions.

#### Vaccination behavior

A logistic regression model was estimated using maximum likelihood estimation was conducted to predict childhood vaccination behavior (see Supplementary File 4 for details). The explanatory power of the model was moderate (Tjur’s R² = 0.25). Of the psychological and social variables, attitudes towards vaccination (OR = 2.12, *p* < 0.001), masculinity (OR = 1.18, *p* = 0.049) and misinformation (OR = 0.67, *p* = 0.002) were significantly associated with vaccination behavior. All other psychological predictors were not related to vaccination behavior (*ps* ≥ 0.107). Regarding demographic variables, children’s age was positively related to vaccination behavior. Compared to children under one year of age, children aged one and over were more likely to have received vaccinations (OR = 1.69-3.57, *ps* < 0.066). Households in Kenya (OR = 9.35, *p* < 0.001) and Malawi (OR = 6.83, *p* < 0.001) were more likely to vaccinate their children than households in Nigeria. Household income showed an inconsistent picture. Compared to households without income, households in the highest income category had a higher probability to vaccinate their children (OR = 4.43, *p* = 0.030). The other income categories did not differ in their vaccination behavior compared to households without income. Moreover, predominantly Muslim households showed lower vaccination behavior (OR = 0.52, *p* = 0.006). Further regressions were conducted to examine the relationship between trust in vaccination information sources, intention to vaccinate and actual vaccination behavior, among other exploratory variables.

### Secondary outputs: Key factors driving vaccination decision-making in SSA Gender of children (gendered norms/gender disparity)

A linear regression model was used to identify the factors associated with beliefs about gender disparity in childhood vaccination (see Supplementary File 5). The model explained a moderate proportion of the variance in gender disparity beliefs (adjusted R² = 0.24, *p* < 0.001). Higher levels of masculinity were significantly associated with stronger beliefs in gender disparity (β = 0.11, *p* < 0.001), indicating that more patriarchal attitudes are linked to a stronger perception that vaccinating girls is more important than vaccinating boys. Similarly, greater endorsement of misinformation (β = 0.23, *p* < 0.001) and higher trust in information sources (β = 0.21, *p* < 0.001) were also associated with stronger gender disparity beliefs. Conversely, more positive attitudes towards vaccination were associated with lower gender disparity beliefs (β = –0.11, *p* < 0.001). Neither religious beliefs nor subjective vaccination knowledge showed significant associations with gender disparity.

Among the demographic variables, caregivers’ level of education and household structure emerged as relevant predictors. Those with a primary education reported stronger gender disparity beliefs than those with no education (β = 0.30, *p* = 0.037). Participants with higher education than primary showed a tendency of stronger gendered norms than those without education (β = 0.24–0.25, *p* > 0.052). Respondents from polygamous households displayed stronger gender disparity beliefs than those from monogamous households (β = 0.21, *p* < 0.001). Household income was negatively associated with gender disparity, suggesting that higher-income households were less likely to hold gendered views on vaccination (β = −0.41 - −0.17, *p* < 0.054).

Cross-country differences were also observed. Compared with Nigeria, caregivers in Kenya and Malawi reported significantly lower gender disparity beliefs (β = –0.48 and β = –0.38, respectively, both *p* < 0.001), reflecting socio-cultural variations in gender norms across the region. As illustrated in Figure 3, these differences were primarily driven by cultural and religious justifications for prioritizing girls in vaccination decisions.

**Figure 3.**
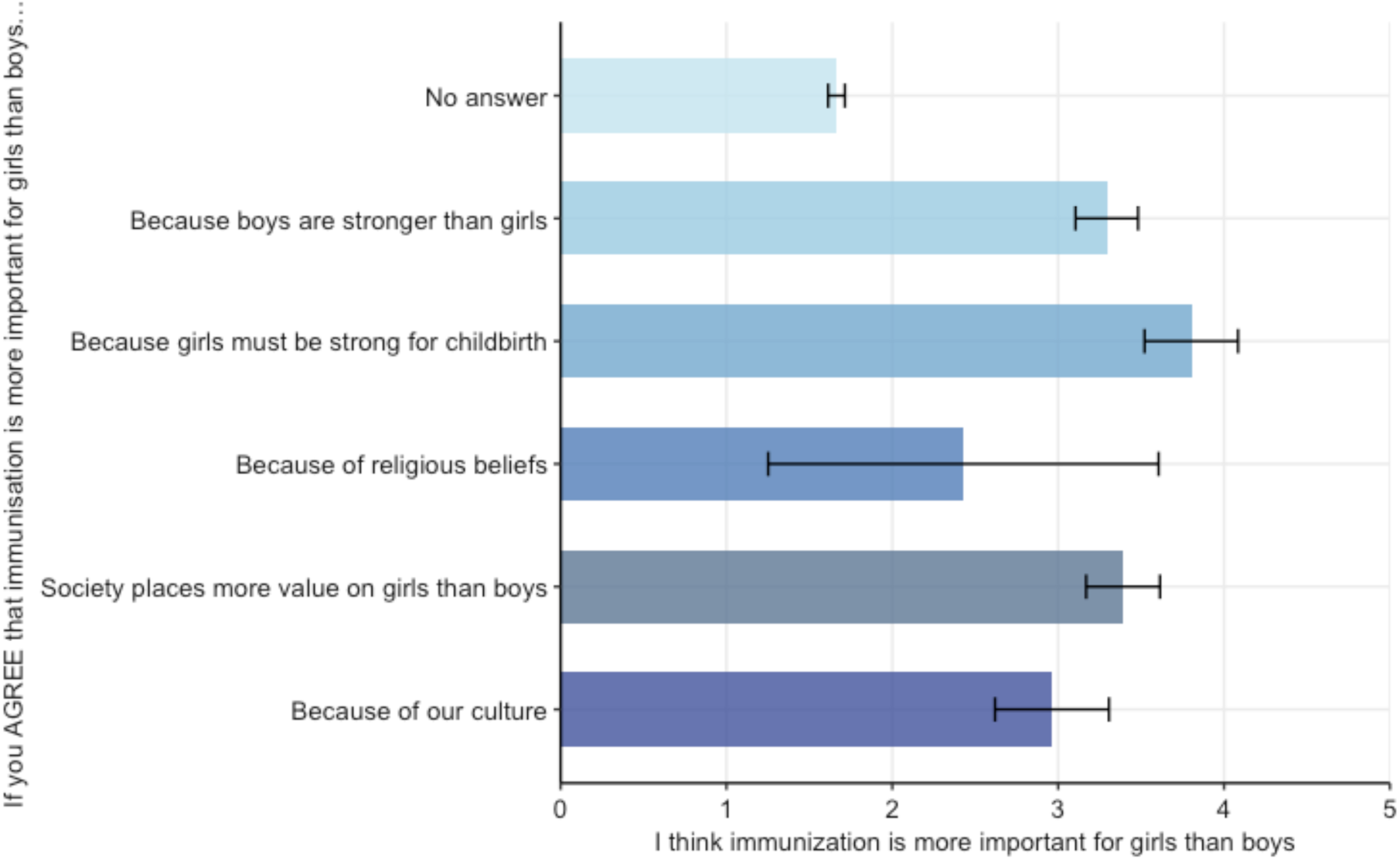
**Socio-cultural and psychological behaviors influencing gender disparity beliefs.** Households with higher disparity perceptions are more likely to rationalize it with patriarchal reasoning. The bars represent the mean values, and the error bars represent the 95% confidence intervals.

### Masculinity

A linear regression model was used to identify the predictors of masculinity, defined as traditional paternal attitudes towards decision-making regarding childhood vaccinations (see Supplementary File 6). The model explained a substantial proportion of the variance in masculinity (adjusted R² = 0.28, *p* < 0.001). Higher gender disparity beliefs (β = 0.10, *p* < 0.001) and greater endorsement of misinformation (β = 0.14, *p* < 0.001) were significantly associated with stronger masculine attitudes. Religious beliefs (β = 0.05, *p* = 0.030) and greater subjective knowledge about vaccination (β = 0.09, *p* < 0.001) were also positively related to masculinity. Trust in information sources and attitudes towards vaccination were not significant predictors. Demographic factors played an important role. Male caregivers reported higher masculinity scores than female caregivers (β = 0.27, *p* < 0.001), and younger caregivers (aged 18 – 24) expressed stronger masculine attitudes than those aged 25 – 34 or 35 – 44 (β = −0.20 and β = - 0.19, *ps* < 0.007). Higher education levels were consistently associated with lower masculinity (βs = −0.68 - −0.32, *ps* < 0.001), indicating that education mitigates traditional gendered beliefs about decision-making in child health. Muslim caregivers reported higher masculinity than Christian caregivers (β = 0.31, *p* < 0.001). At the household level, respondents from polygamous households exhibited higher masculinity scores than those from monogamous households (β = 0.24, *p* < 0.001). Household income showed mixed associations; only caregivers in the lowest income bracket reported significantly lower masculinity than those with no income (β = −0.17, *p* = 0.032). Cross-country differences between Nigeria, Kenya, and Malawi were also evident, aligning with previous findings (β = −0.20 and β = −0.57, *ps* < 0.007).

### Misinformation

A linear regression model was used to analyze predictors relating to beliefs relating about misinformation concerning vaccination (see Supplementary File 7). The model explained a substantial proportion of variance (adj. R² = 0.34, *p* < 0.001). Misinformation was positively associated with gender disparity (β = 0.20, *p* < 0.001), masculinity (β = 0.13, *p* < 0.001) and religious beliefs (β = 0.15, *p* < 0.001). These findings suggest that adherence to traditional gender norms and religiosity are associated with a greater tendency to endorse misinformation. Conversely, trust in information sources (β = –0.06, *p* = 0.008) and positive attitudes towards vaccination (β = –0.24, *p* < 0.001) were negatively related to misinformation. This suggests that confidence in reliable information and favorable perceptions of vaccination reduce susceptibility to misinformation. Educational level also emerged as a significant factor. Caregivers with secondary (β = –0.27, *p* = 0.022) or tertiary education (β = –0.30, *p* = 0.013) reported lower levels of misinformation than those with no formal education. The age and gender of caregivers were not significantly related to misinformation (*ps* ≥ 0.059). Belief in misinformation was positively associated with the age of the child (β > 0.11, *p* < 0.053). Parents of male children (β = 0.14, *p* < 0.001) or children from diverse backgrounds (β = 0.48, *p* = 0.009) reported higher levels of misinformation than parents of female children. Household income showed inconsistent associations; however, higher-income households were less likely to endorse misinformation (β = –0.22, *p* = 0.020) and households without income. Finally, country-level differences were observed. Compared to Nigeria, misinformation beliefs were lower in Malawi (β = –0.50, *p* < 0.001), but slightly higher in Kenya (β = 0.11, *p* = 0.045).

### Contexts

#### Country-level differences within examined variables

To explore variations at the country level, regression models were estimated with Nigeria as the reference group (see Table 1). Significant cross-national differences emerged in both psychosocial predictors and vaccination outcomes. Compared to Nigeria, caregivers in Kenya reported remarkably lower levels of gender disparity (β = –0.42, *p* < 0.001) and masculinity (β = –0.40, *p* < 0.001), as well as higher vaccination intentions (β = 0.20, *p* < 0.001) and a greater likelihood of vaccinating their children (OR = 3.64, *p* < 0.001). This pattern suggests that less pronounced gendered norms in Kenya are associated with stronger vaccine acceptance and uptake. In Malawi, gender disparity (β = –0.82, *p* < 0.001), misinformation (β = –0.80, *p* < 0.001) and masculinity (β = –0.79, *p* < 0.001) were all substantially lower than in Nigeria. Despite a somewhat lower vaccination intentions (β = 0.66, *p* < 0.001), caregivers in Malawi displayed the highest probability of actual vaccination behavior (OR = 13.13, *p* < 0.001).

**Table 1:** Cross-country variation of variables.

| *Predictors* | Gender Disparity |  | Misinformation |  | Masculinity |  | Intention to vaccinate (child) |  | Vaccination behavior (child) |  |
| --- | --- | --- | --- | --- | --- | --- | --- | --- | --- | --- |
| | $\beta$ | *p* | $\beta$ | *p* | $\beta$ | *p* | $\beta$ | *p* | OR | *p* |
| (Intercept) | 0.32<br>(0.27 – 0.38) | <0.001 | 0.15<br>(0.09 – 0.21) | <0.001 | 0.31<br>(0.25 – 0.37) | <0.001 | -0.20<br>(-0.26 – -0.14) | <0.001 | 3.44<br>(2.97 – 4.00) | <0.001 |
| Kenya vs. Nigeria | -0.42<br>(-0.51 – -0.34) | <0.001 | 0.00<br>(-0.08 – 0.09) | 0.915 | -0.40<br>(-0.48 – -0.31) | <0.001 | 0.20<br>(0.12 – 0.29) | <0.001 | 3.64<br>(2.76 – 4.84) | <0.001 |
| Malawi vs. Nigeria | -0.82<br>(-0.92 – -0.71) | <0.001 | -0.80<br>(-0.90 – -0.69) | <0.001 | -0.79<br>(-0.90 – -0.69) | <0.001 | 0.66<br>(0.55 – 0.77) | <0.001 | 13.13<br>(7.27 – 26.73) | <0.001 |
| Observations | 2474 |  | 2448 |  | 2470 |  | 2469 |  | 2457 |  |
| R<sup>2</sup> / R<sup>2</sup> adjusted | 0.092 / 0.091 |  | 0.098 / 0.097 |  | 0.086 / 0.085 |  | 0.056 / 0.055 |  | 0.065 |  |

*Note.* Gender disparity was measured using a scale where higher values indicated higher gender equality. The factor ‘country’ used Nigeria as the base level. $\beta$ = standardized regression coefficient, OR = Odds ratio, and the values in parentheses correspond to 95% confidence intervals.

Overall, these findings highlight meaningful regional variations across SSA. Kenya and Malawi exhibit lower endorsement of patriarchal norms and misinformation, which correspond to higher vaccination outcomes relative to Nigeria. However, the discrepancy between intention and actual behavior in Malawi suggests that contextual factors beyond individual beliefs also play an important role in shaping vaccination practices.

## DISCUSSION

This study improves our understanding of how caregivers make decisions about vaccination in SSA by integrating psychological, social and cultural determinants into a single explanatory model. In summary, the findings confirm that psychosocial and normative factors, particularly misinformation, masculinity, and gender norms and disparity, play a pivotal role in shaping vaccination decision-making among caregivers in SSA. The findings revealed that beliefs in misinformation, masculinity, gender disparity, and religious doctrines consistently undermine vaccination attitudes, intentions, and behaviors across the three SSA countries assessed. Conversely, trust in credible information sources and positive attitudes towards vaccination emerged as key facilitators. These insights underscore the complex interplay between gender norms, informational environments, and socio-economic realities shaping vaccination behavior. The study outcomes confirmed the hypotheses based on TPB assumptions that caregivers’ vaccination decision-making is patterned rather than random, as shown in Figure 4.

**Figure 4:**
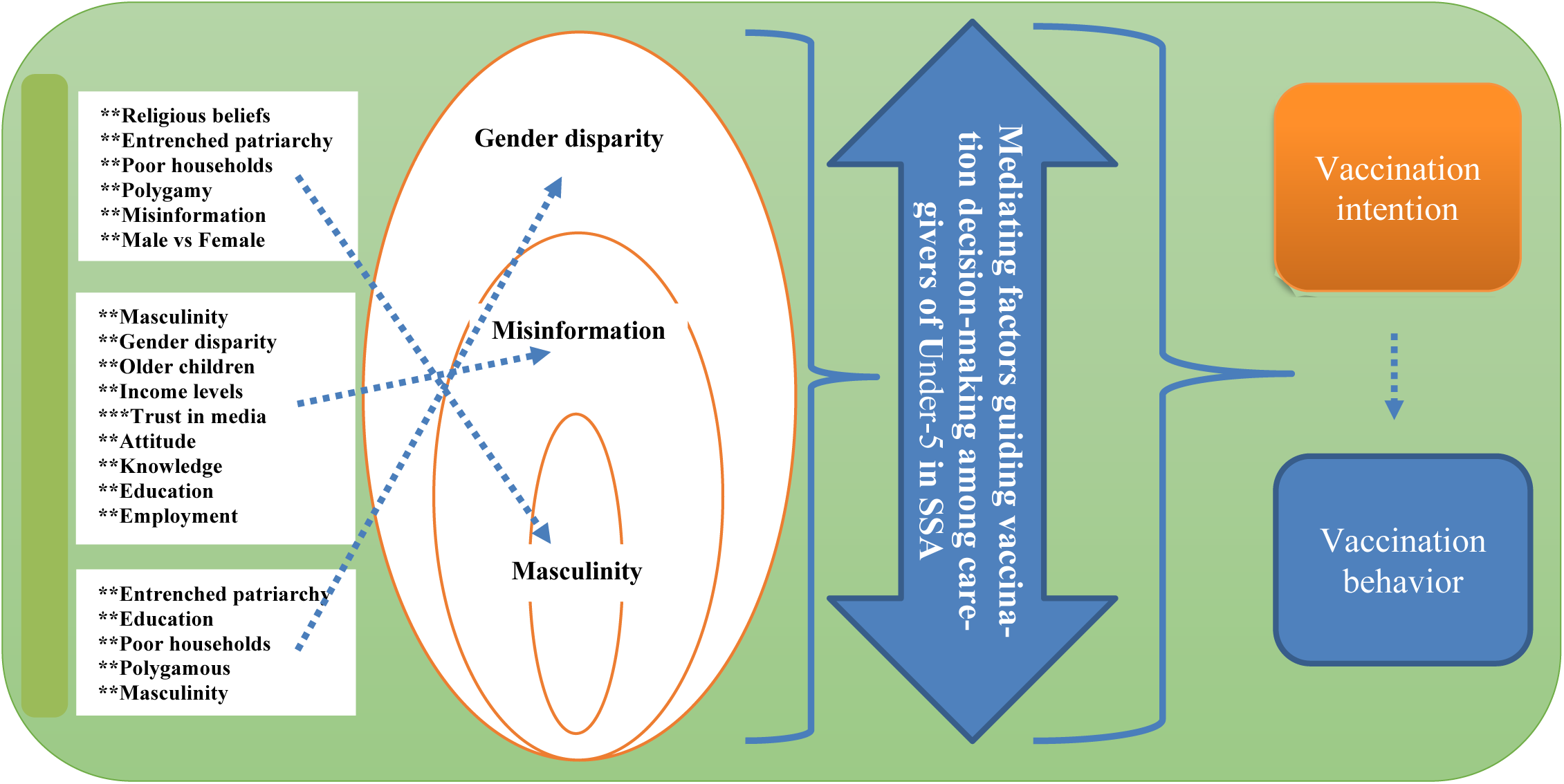
**Framework of the key measures of vaccination intention and behavior in SSA.**

The association between masculinity and gender disparity is consistent with previous findings that patriarchal decision-making structures restrict women’s autonomy in child health matters [15,16,20]. However, the present study builds on existing research by showing that masculinity not only restricts maternal decision-making space, but also directly correlates with lower vaccination intentions among fathers. This highlights the importance of gender-transformative public health strategies that encourage men to play an active role in child health rather than acting as gatekeepers. Similarly, misinformation was negatively associated with both intention and behavior, reaffirming global evidence on the detrimental effects of exposure to vaccine myths. Nevertheless, the observed link between misinformation and gendered beliefs suggests that misinformation may be socially embedded within prevailing gender ideologies, rather than being purely informational. These dynamics may explain why standard awareness campaigns often have limited success in highly patriarchal contexts.

Cross-country differences also reveal the moderating role of social context. Kenya and Malawi, with comparatively lower masculinity and misinformation tendencies, had higher vaccination uptake than Nigeria, emphasizing how structural gender equality and supportive public information ecosystems can inspire vaccine acceptance.

### Childhood vaccination intention and behavior in SSA

One unique outcome of this study is its highlighting of the central role of fathers in shaping childhood vaccination decisions in SSA. The study shows that children are more likely to be vaccinated when their fathers have positive attitudes and that, as illustrated in Figure 5 and supported by previous studies [38–40], boys consistently have better vaccination outcomes than girls. These patterns reflect the strong influence of masculinity, gender preference and patriarchal norms, which are also closely linked to the persistence of myths and misinformation. In addition, religious beliefs, particularly within Muslim households and some Christian sects, remain important drivers of vaccine hesitancy, reinforcing the earlier evidence that belief systems strongly influence perceptions of, and demand for, vaccines [17,20,30,41]. Therefore, culturally and religiously sensitive interventions are needed. The study also show that masculinity, gender disparity and misinformation jointly undermine vaccination intention and behavior in various settings, with notable sub-regional variations. For example, Nigeria has a lower childhood vaccine uptake than Kenya and Malawi, with Muslim households and lower-income groups disproportionately affected [42,43]. This study advances our understanding by demonstrating that caregivers’ vaccination behavior in SSA is shaped by intersecting factors, requiring multi-layered, context-specific intervention strategies.

**Figure 5.**
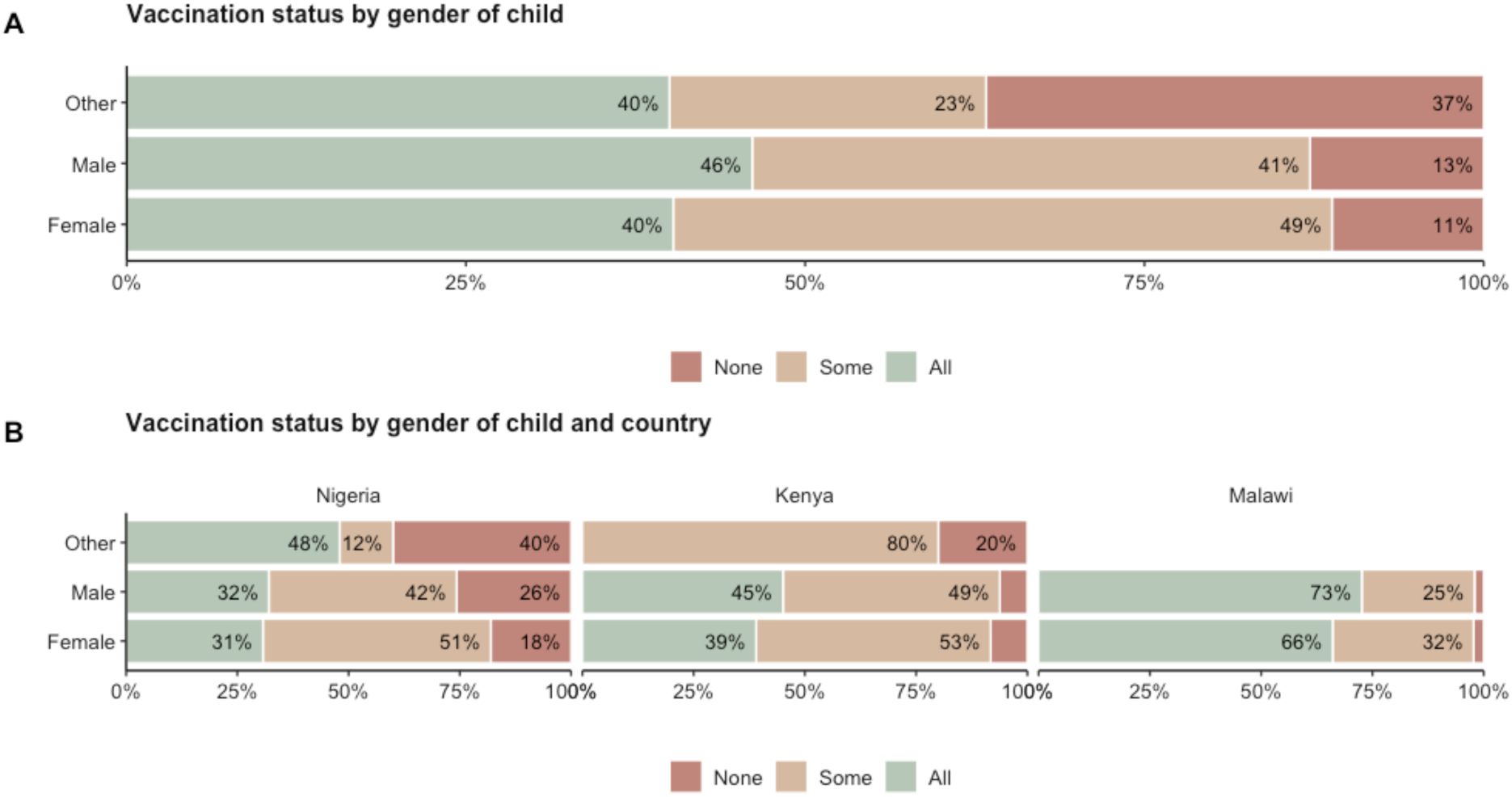
**Vaccination status of child**. Panel A shows the vaccination status distribution based on gender of the child. Panel B shows the vaccination status of the child stratified by child’s gender and country.

### Gender of child (gender disparity)

As shown in Figure 6, this study provides novel evidence that positive attitudes towards gender disparity are inversely related to attitudes towards vaccination. This suggests that support for gender-neutral vaccination is associated with better childhood vaccination behavior, and vice versa. Interestingly, some caregivers perceived vaccinating girls as more important vaccinating boys, despite evidence that boys generally have higher vaccination coverage. This apparent contradiction aligns with earlier studies [38,43] and reflects deeply rooted gender stereotypes of “strong” versus “weak” children, rather than concerns about actual inequalities in vaccination favoring boys, as shown in Figure 5. Figure 3 further illustrates how beliefs such as “girls must be strong for childbirth” and “boys are stronger than girls” underpin these perceptions, highlighting how socio-cultural norms shape vaccination decision-making processes [15,16,38,44]. The stronger the gendered norms, the greater their influence on vaccination decision-making.

**Figure 6:**
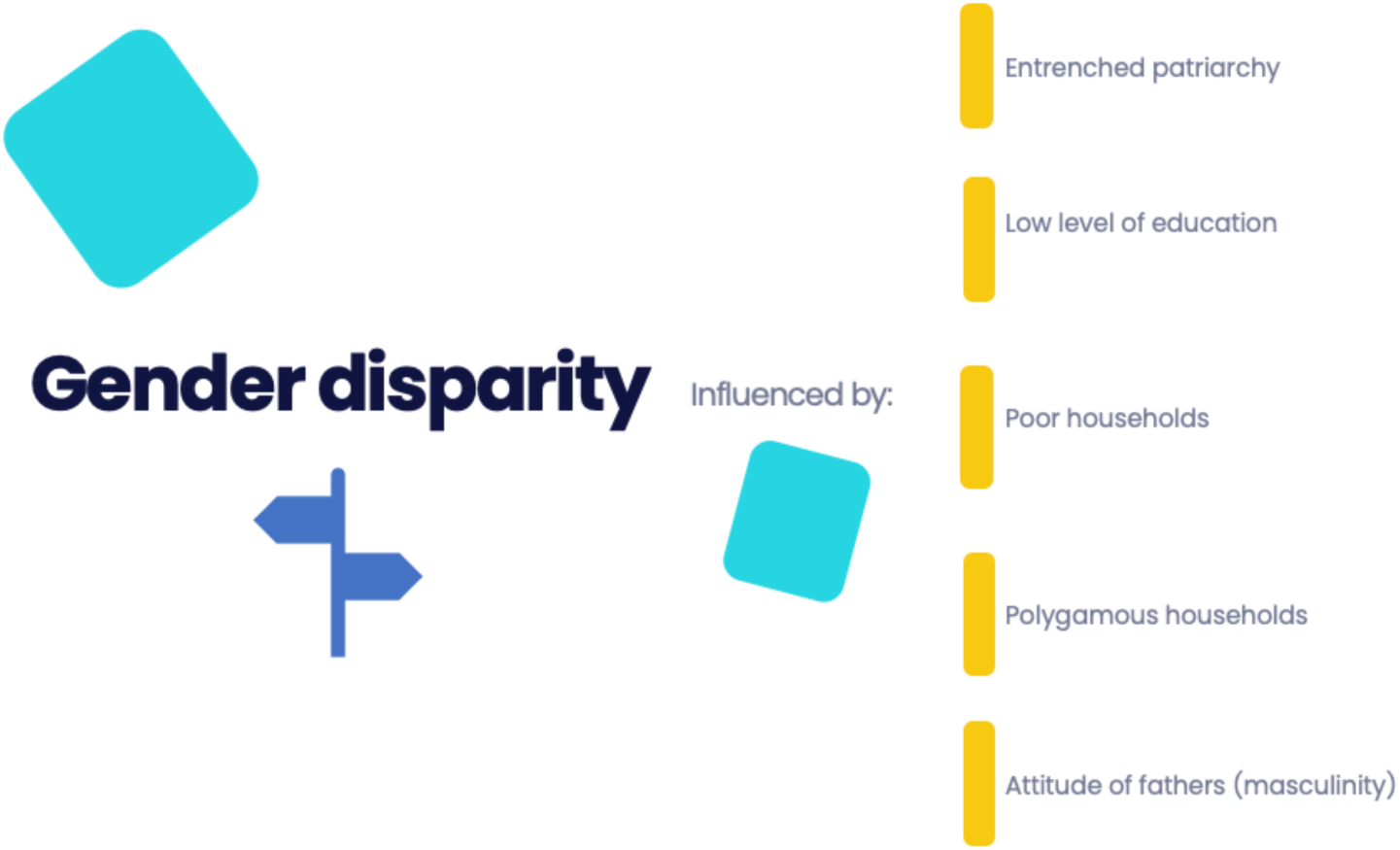
**Gender disparity influencers.**

It is novelle to find that beliefs in gender disparity are more prevalent in polygamous, poorer and more religious households, particularly among Muslim families. These beliefs are associated with lower income and education levels. In Nigeria, despite minimal gender differences in coverage, as shown in Figure 5, more favorable attitudes towards vaccinating girls were expressed by more educated and polygamous households, suggesting shifting norms [9,45,46]. This is because polygamy, entrenched patriarchy and lower levels of education are associated with northern Nigeria, where vaccination coverage is lower (below 40% on average) than in the south (above 70% on average); which is more educated, liberal and has a lower prevalence of polygamous households [9,45,46]. Thus, caregivers’ vaccination behavior in SSA is shaped by complex and sometimes contradictory interactions between gender norms, household structure (e.g., polygamy), religious beliefs, education, and socioeconomic status.

### Masculinity (attitude of fathers)

It is not unexpected that masculinity is strongly linked to beliefs in gender disparity and plays a central role in caregivers’ vaccination decision-making in SSA. In highly patriarchal households, where fathers dominate decision-making, preferences for protecting one gender over another are shaped by attitudes, religious beliefs, and beliefs in misinformation [17,20,30,47]. Therefore, the notion that girls should be prioritized for vaccination reflects the “weaker versus stronger” gender stereotype [48,49], while also aligning positively with religious beliefs and misinformation, as shown in Figure 7. These interactions highlight how traditional masculine identities shape interpretations of vaccine information and trust [50,51]. The disappearance of the negative association between masculinity and vaccination attitudes after demographic adjustment confirms that gender norms are often mediated by socio-demographic factors [52,53]. Consistent with prior studies [15,54–58], restrictive gender norms that limit women’s autonomy remain a major barrier to childhood vaccination. Higher masculinity attitudes were more prevalent among younger, male, less educated, Muslim caregivers and those in polygamous households [59,60], particularly in Nigeria, where low coverage is concentrated in the conservative north. This illustrates how environmental factors can influence vaccination decision-making.

**Figure 7:**
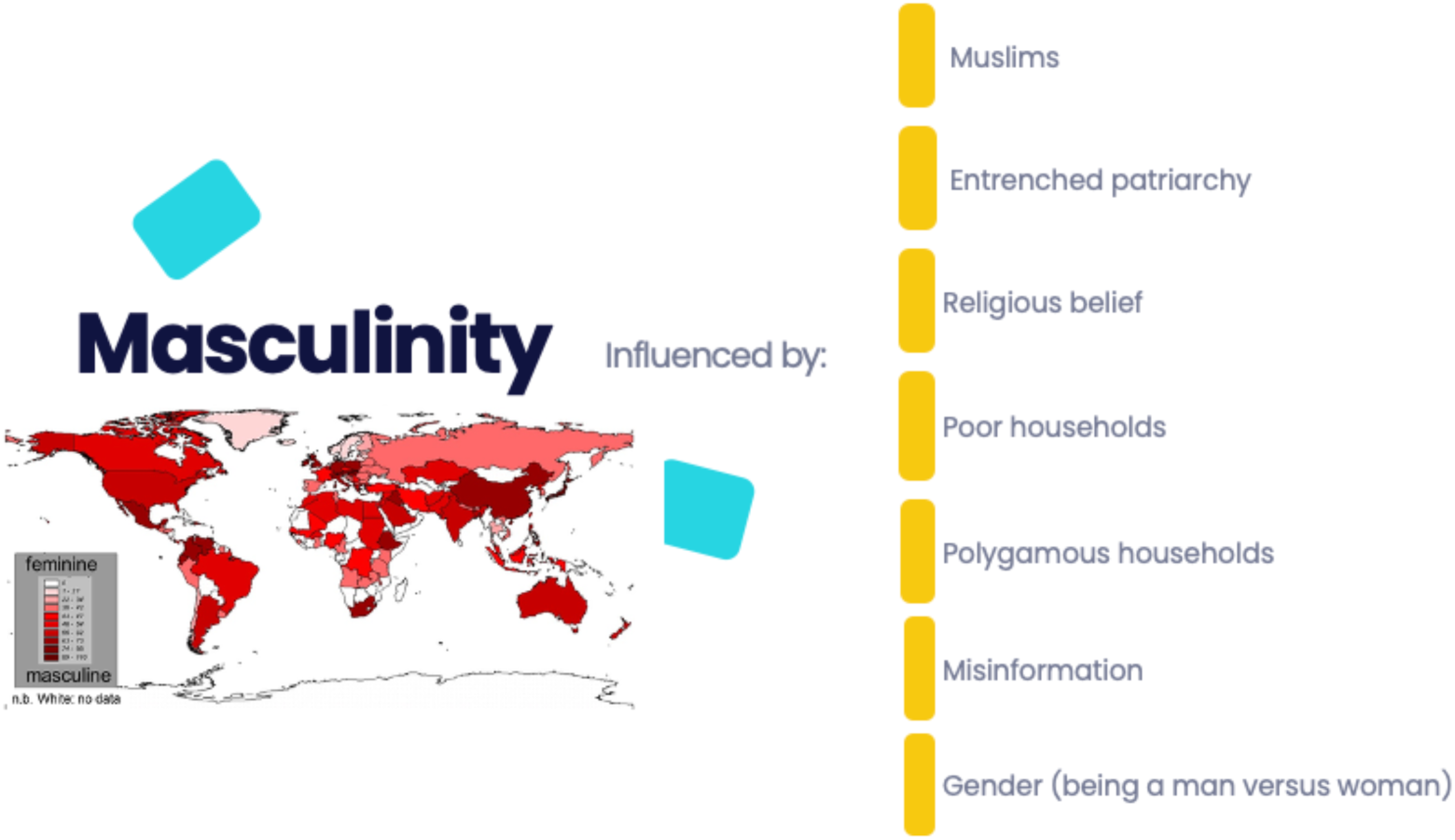
**Masculinity belief enablers**

In several African cultures, the role of fathers is often defined by traditional norms and expectations, which can vary significantly across sub-regions and socio-economic contexts [61,62]. However, understanding these dynamics is crucial for contextualizing the household decision-making processes, including those relating to vaccination. Interventions to improve vaccination uptake in SSA should consider the position of fathers within households and how scientific knowledge about the impact of childhood vaccinations on community well-being is disseminated, taking into account socio-cultural expectations. Indeed, given the increasing evidence of very low vaccination coverage in certain SSA settings, interventions that specifically target men should be prioritized to increase demand for vaccination.

The central role of masculinity therefore highlights how fathers’ beliefs and authority can either facilitate or hinder childhood vaccination, positioning men as critical enabler or barrier in vaccination decision-making process. This study demonstrates that masculinity, religious beliefs, misinformation, and socio-economic disadvantage are interconnected drivers of low vaccination uptake in SSA. This underscores the need for interventions that engage fathers directly and promote gender-neutral vaccination messages.

### Misinformation

This study shows that susceptibility to misinformation is socially patterned rather than random. It is also strongly shaped by patriarchy, masculinity and religious beliefs, particularly in Muslims households. However, vaccine hesitancy has also been observed among Christians, particularly in Malawi [16]. As Figure 8 shows, tackling misinformation requires an encompassing approach, as it is uniquely associated with masculinity and gender disparity. This indicates that vaccine myths are reinforced by gender norms and hierarchical social structures. This includes the fact that fathers’ attitudes and beliefs about protecting the “weaker” gender, as well as culturally grounded myths, interact to shape vaccination decision-making, particularly in patriarchal contexts, as observed in Kenya, Nigeria and Malawi. The strong link between misinformation, masculinity, and religion is consistent with existing evidence that traditional gender norms and certain religious frameworks foster skepticism towards vaccination [63–66]. Conversely, households with stronger beliefs in gender disparity are also more likely to accept misinformation, suggesting that vaccine information is processed through a gendered lens. Therefore, religious beliefs fuelled by myths and mis-or-disinformation still play a dominant role in vaccination decision-making in SSA.

**Figure 8:**
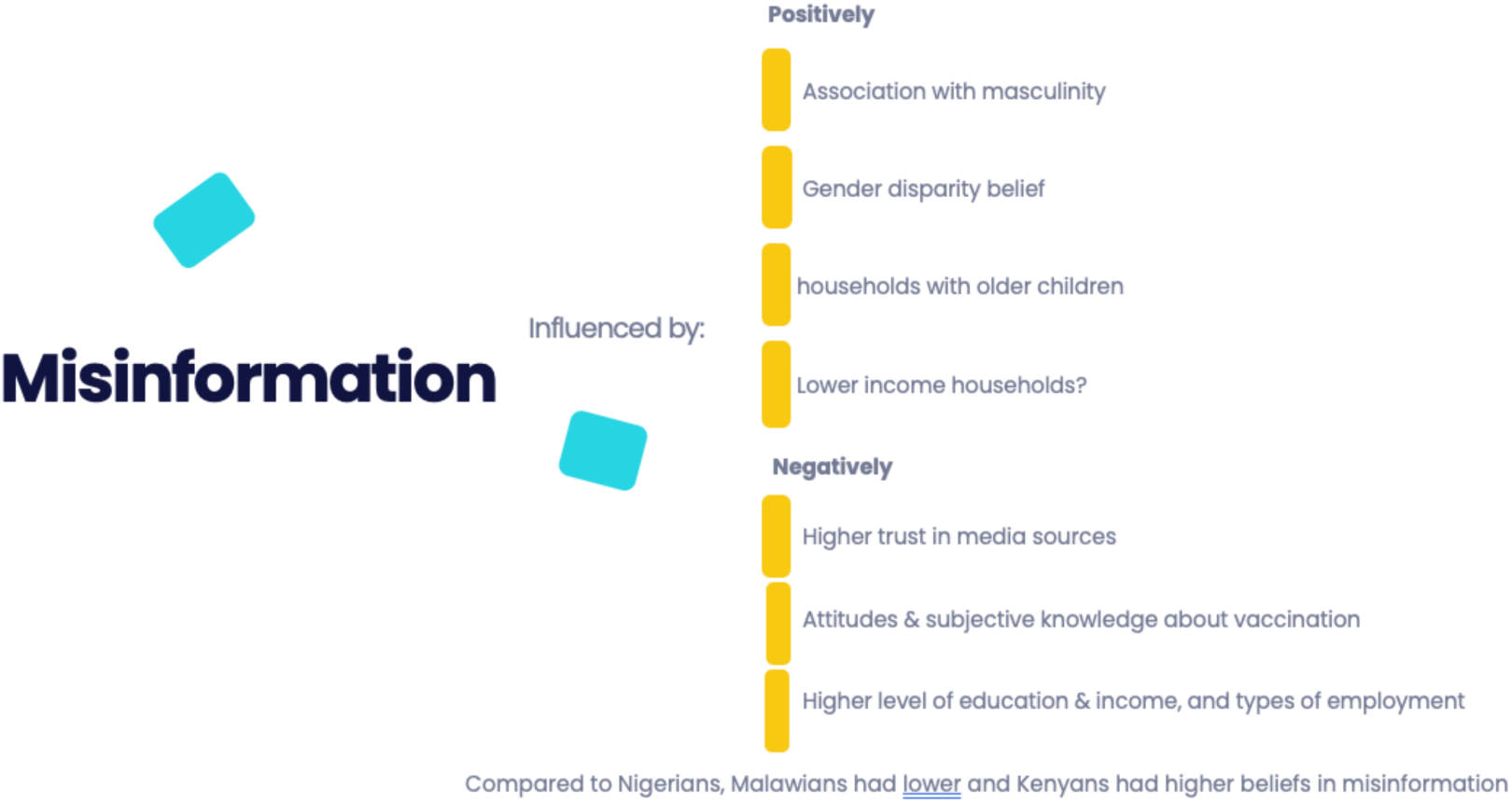
**Factors driving misinformation in the study settings**

The impact and patterns of misinformation should be considered alongside country-level differences. For example, caregivers in Malawi are less susceptible to misinformation than those in Nigeria and Kenya, which partly explains why Malawi has comparatively higher vaccination coverage. While education generally reduces vulnerability to misinformation, recent evidence [16,67,68] supports the findings of this study that hesitancy can also be high among the educated and wealthy, underscoring the need for tailored communication strategies. It is also interesting that households with older children, children over one year old, the unemployed, and those with a low income are more likely to believe vaccine misinformation. Therefore, vaccine communication strategies should be tailored to specific socio-economic and household contexts, rather than treating misinformation as a uniform problem. In addition, trust in information sources is paramount, as caregivers who rely on HCWs exhibit higher vaccination intentions than those who depend on social media or informal networks. This highlights the need to strengthen the visibility, training, and communication capacity of HCWs. Interventions should integrate a gender-sensitive approach, religious engagement, misinformation management and HCW-led communication to effectively improve vaccination uptake in SSA.

Importantly, this study provides practical guidance on interventions. Debunking myths and mis or disinformation requires a structured, evidence-based approach, such as the “sandwich model” (see Figure 9) [38,69,70], which begins and ends with “*facts”* while explicitly addressing and correcting misinformation. Although the design was originally intended for patient consultations, it is also suitable for interventions targeting the debunking of misinformation, especially in limited-resource settings such as SSA, where the population has a high level of trust in HCWs. Interventions using this model should start with the *‘fact’* or truth about vaccine safety and effectiveness. Secondly, ‘*myth’* and misinformation about vaccines and vaccination need to be discussed. E.g., the perception that childhood vaccines contain family planning antigens designed to damage the reproductive ability of the African population. Thirdly, the *‘fallacy’* against vaccines should be explained in detail using scientific evidence to show why the misinformation or myths are false. Fourthly, the *‘damage done’* by the myth or misinformation should be explained using context-specific illustrations, as these are a catalyst for vaccine hesitancy. Lastly, interventions targeting myths and mis or dis-information should consistently reinforce the *‘fact’*, ensuring it remains the primary information that caregivers take away about vaccines and vaccination.

**Figure 9:**
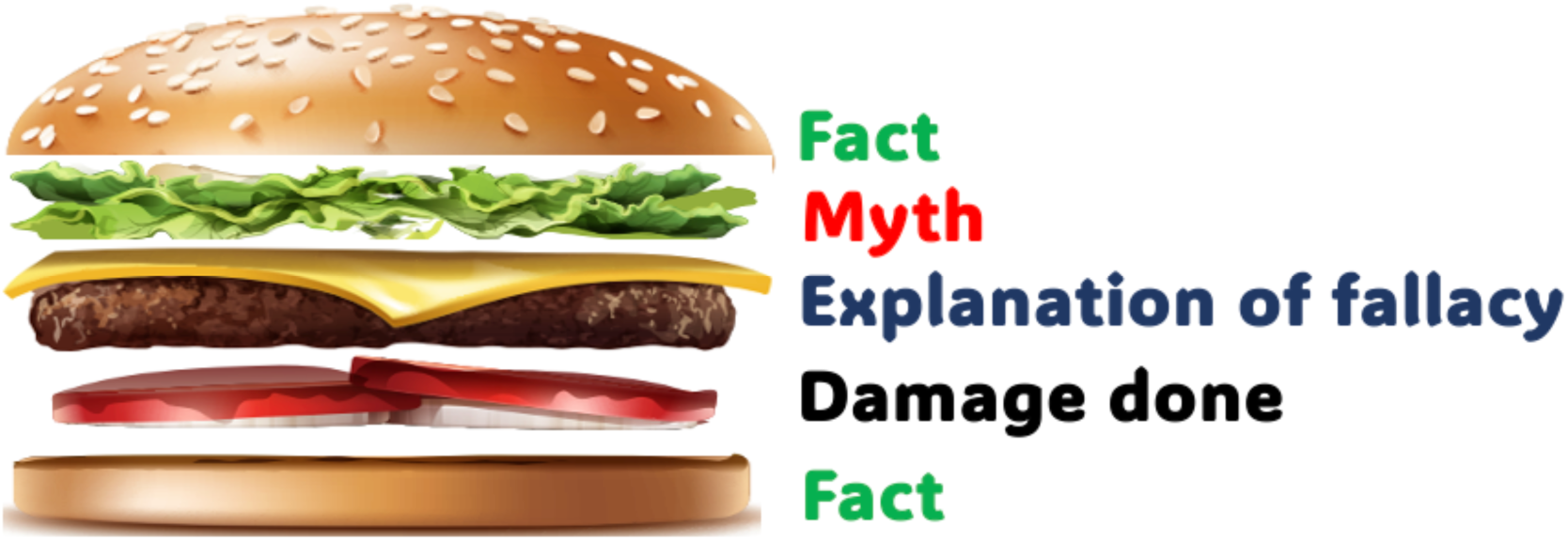
**The sandwich model for tackling vaccine myths and misinformation [38].**

### Summary implication of other outputs

This body of evidence underscores that childhood vaccination intention and behavior in sub-Saharan Africa (SSA) are shaped by a complex interaction. A key factor in improving uptake is strengthening caregivers’ trust in vaccines and in credible information sources, particularly when communication is transparent and delivered by trusted messengers, such as HCWs, as found in this study. Although perceived knowledge alone shows a weaker direct link to vaccination behavior, nevertheless, reinforcing accurate knowledge of vaccinations positively influences attitudes and intentions. Importantly, the study shows that vaccination decisions are also socially embedded: with peer influence and community norms strongly shaping caregivers’ decision. This indicating that community-based and peer-led approaches can effectively normalize pro-vaccination attitudes. Therefore, this study reinforces the idea that vaccination demand cannot be addressed through information at individual level alone but requires interventions that engage social networks and collective expectations. This suggests that structural or programmatic factors, such as more effective immunization campaigns or stronger community health engagement, could lead to high vaccination uptake even where intention scores are low.

The outcome of this study further demonstrates that structural and relational factors critically shape vaccination behavior. Male caregivers, who are less likely to vaccinate their children, are strongly influenced by patriarchal norms and beliefs in gender disparity, highlighting the need for gender-transformative interventions that actively engage men and challenge restrictive norms. Household income and structure also matter: economic vulnerability amplifies misinformation and gender inequality, while polygamous households present complex decision-making dynamics that require tailored engagement. At the same time, empowering young and educated women can enhance vaccination uptake by strengthening household health decision-making, although interventions must remain culturally sensitive to avoid reinforcing inequalities. Overall, strengthening the role of HCWs as trusted messengers through training, support and visibility within the Expanded Program on Immunization remains critical for countering misinformation, building trust, and increasing equitable vaccination demand across SSA.

This study is not without some limitations. Firstly, the cross-sectional design may limit the ability to draw firm conclusion about causation, since the relationships between masculinity, gender disparity, misinformation, intention and vaccination decision-making were observed at a single point in time and may be influenced by unmeasured confounding factors. Secondly, the reliance on self-reported data may introduce recall and social desirability bias, particularly regarding sensitive issues such as gender norms, religious beliefs, and vaccination practices, where respondents may provide socially desirable responses. Thirdly, although the study included multiple countries, the findings may not be fully generalizable to all SSA contexts due to differences in culture, politics and health systems across countries and within sub-national regions. Finally, the study primarily focused on demand-side factors and did not comprehensively assess supply-side constraints, which may interact with behavioral drivers. Despite these limitations, the scientific rigor is of a sufficiently high standard to support the reported conclusions. Furthermore, the study provides a robust basis for future longitudinal and mixed-methods research.

## CONCLUSION

This study provides strong empirical evidence that the vaccination intentions and behaviors of caregivers in SSA are shaped by the core TPB constructs of attitudes, subjective norms, and perceived behavioral control. Caregivers’ attitudes were reflected in their perceptions of vaccine safety and usefulness, which were strongly influenced by misinformation, trust in information sources, and beliefs about masculinity and gender disparity. Subjective norms were evident in the significant influence of fathers, peer influence, religious beliefs and community expectations. This demonstrates that vaccination decision-making is deeply embedded in social approval and conformity to dominant norms. Perceived behavioral control was captured through factors such as household income and structure (e.g., polygamy), access to reliable information, and trust in HCWs, all of which shaped caregivers’ ability and confidence to act on positive intentions.

The study also shows that, when attitudes are positive, norms are supportive, and trusted health information is accessible, vaccination intention translates more strongly into actual behavior. Therefore, improving vaccination uptake in SSA requires interventions that simultaneously transform attitudes (by countering misinformation), reshape social norms (by engaging fathers, religious leaders, and communities), and strengthen perceived behavior control (by empowering caregivers with trusted support systems).

Improving vaccination uptake requires more than improving supply or access; it demands gender-sensitive, trust-centered, and socially grounded strategies. Public health interventions must actively engage fathers, challenge harmful beliefs about masculinity and gender disparity, and position HCWs as central, trusted communicators of vaccine information. At the same time, policies should harness peer influence, community networks, and social mobilization to counter misinformation and reinforce pro-vaccination norms. It is essential to empower HCWs with communication tools, integrate culturally sensitive messaging and design inclusive approaches that consider household structure, income and gender dynamics. Overall, strengthening equitable child health outcomes and vaccination uptake across the SSA region will depend on integrated strategies that address misinformation, gender disparity beliefs, and masculinity tendencies simultaneously.

## DECLARATIONS

### Ethics approval and consent to participate

The study was conducted in accordance with the principles of the Declaration of Helsinki and approved by the relevant Health Research Ethics Committee (reference number FHREC/2023/01/48/04-03-23). Informed consent was obtained from all participants.

### Consent for publication

All authors approved the final manuscript.

### Availability of data and materials

The datasets used and/or analyzed during the current study are available at https://osf.io/sa4vp/

## Author’s Contribution

Conceptualization: GCA Methodology: GCA Investigation: GCA.

Result Analysis: GCA and LK

Writing – original draft: GCA

Writing – review & editing: GCA and LK

## Data Availability

The datasets used and/or analyzed during the current study are available at https://osf.io/sa4vp/

## ACKNOWLEDGMENTS

We would like to express our sincere gratitude to all the participants in this study who generously agreed to be interviewed despite their busy schedules. We would also like to acknowledge the field workers who provided support during the data collection process.

